# Reliability of Extraocular Muscle Measurements on Magnetic Resonance Imaging in Graves’ Disease: A two-rater case study

**DOI:** 10.1101/2025.09.19.25336132

**Authors:** Rafael Luccas, Fábio Mota Gonzalez, Cristiane Abbehusen Lima Castelo Branco, Cinthia Minatel Riguetto, Denise Engelbrecht Zantut-Wittmann, Fabiano Reis

## Abstract

Thyroid eye disease (TED) is based mainly on clinical and ophthalmological findings, but imaging exams such as magnetic resonance imaging (MRI) is helpful on its identification and follow-up when the disease is clinically active. The main characteristic of TED is the enlargement of extraocular muscles that can be easily assessed with good quality of imaging but it is still challenging to evaluate it in the scenario of symmetric mild disease, where the patient-specific normal measure parameter is lost. Considering these challenges, the objective of this study was to evaluate two different raters with similar clinical experiences on measuring MR images to obtain metrics of extraocular musculature and investigate the method reliability isolated from the clinical perspective. A total of 31 MR images were collected between 2019 and 2021. The images are from 3 groups: Patients with Graves’ Disease (GD) and active inflammatory ophthalmopathy, patients with GD and inactive ophthalmopathy and healthy control group without evidence of autoimmune thyroid disease. The radiologists measured the inferior rectus, superior rectus, medial rectus, lateral rectus, superior oblique and inferior oblique muscle thickness, as well the proptosis divided by eye. They also identified the increase of extraconal fat, intraconal fat and changes in lacrimal gland. Both raters were blinded to each other measurements and to the patient group. On an overview, we could identify differences in the results for each rater. One rater had more reliable results evaluating eye muscles, while the other had better results assessing other eye structures, which can be due to the experience with the disease and with the imaging analysis. We concluded that the method is not reliable as a standalone evaluation while there is no gold standard on how to perform it, and must continue to be used as supplementary information to corroborate clinical and laboratorial findings.

## Introduction

Thyroid eye disease (TED) affects up to 50% of the patients with Graves’ hyperthyroidism and the symptoms can include from mild ocular irritation to vision loss (1, 2). Despite TED diagnosis being based mainly on clinical and ophthalmological findings, imaging exams such as computed tomography (CT) and magnetic resonance imaging (MRI) are helpful on its identification and follow-up when the disease is clinically active (3). The isolated clinical evaluation of the disease can lead to misdiagnosis considering that several TED symptoms can also be present on other ophthalmopathies, leading to a possible error of the adequate patient treatment.

There has been developed some studies to determine normative data that defines which measures are considered normal for extraocular muscles establishing an average value for each muscle (4–7), but it is not considered a norm because it presents variations and overlap cases on diseased and healthy muscles. When the disease affects mainly the extraocular muscles it is considered myogenic, while when it affects only adipose tissue, it is considered lipogenic (8).

The most frequently involved muscle in clinical myopathy is the inferior rectus muscle, followed respectively by medial, superior and lateral rectus muscles. Multiple muscles can be involved bilaterally and/or simultaneously in 76% to 90% of the cases and extraocular muscle enlargement is the most common sign usually affecting older and younger patients differently (9).

Depending on the most enlarged muscle size and position, it can cause proptosis, however muscle enlargement analysis is not precise or sensitive and should be used as adjuvant evidence for diagnosis (10). Proptosis is usually measured by a Hertel exophthalmometer during clinical evaluations but imaging techniques can also be used and it is graded from 1 to 3, where grade 1 is when more than two-thirds of the anteroposterior diameter of the eyeball is projected to the front of the line; grade 2 is when the posterior pole of the globe border the line; and grade 3 is when the entire eyeball is projected in front of this line (11).

Exophthalmos in the transverse section and extraocular muscle involvement in the coronal section are commonly regarded as diagnostic criteria through radiology evaluation. The measurement of exophthalmos is relatively complex and prone to error (The vertical distance from the front of the eye to the inter-zygomatic line is necessary) for being researcher dependent (12, 13). Some other factors that can lead to the exophthalmos are the retro-orbital fat expansion and lacrimal gland prolapse, which combined with other symptoms can result in irritation, ulceration and the risk of eventual visual loss (14).

The objective of this study was to evaluate two different raters with similar and large clinical experiences on measuring MR images to obtain metrics of extraocular musculature and investigate the method reliability isolated from the clinical perspective.

## Method

A total of 31 MR images were collected at State University of Campinas Hospital between 2019 and 2021. The images came from 3 groups of individuals: Patients with GD with active inflammatory ophthalmopathy, patients with GD and inactive ophthalmopathy and healthy control group. To participate in the study, patients had to sign a paper informed consent form after reading, understanding and clarifying any potential doubts regarding the study, consenting in free will on the participation. Patients also had to be above 18 years old and have a BMI (Body Mass Index) lower than 30 kg/m². The images were obtained using a Philips Achieva 3T, 32 directions with a b-value of 900s/mm² and analyzed using the RadiAnt DICOM Viewer, 64-bit version.

The raters, two radiologists with over 20 years of experience in MR imaging in head and neck, measured the inferior rectus, superior rectus, medial rectus, lateral rectus, superior oblique and inferior oblique muscles thickness (Figure 1), as well the proptosis divided by eye. They also identified if there was an increase of extraconal fat, intraconal fat and changes in lacrimal gland. Both raters were blinded to each other measurements and to the patient group. The measurement technique used to measure muscle involvement was described by Xu, 2017 (15), using the largest diameter of the middle section of each muscle, as exemplified on Figure 1.

**Fig 1.**
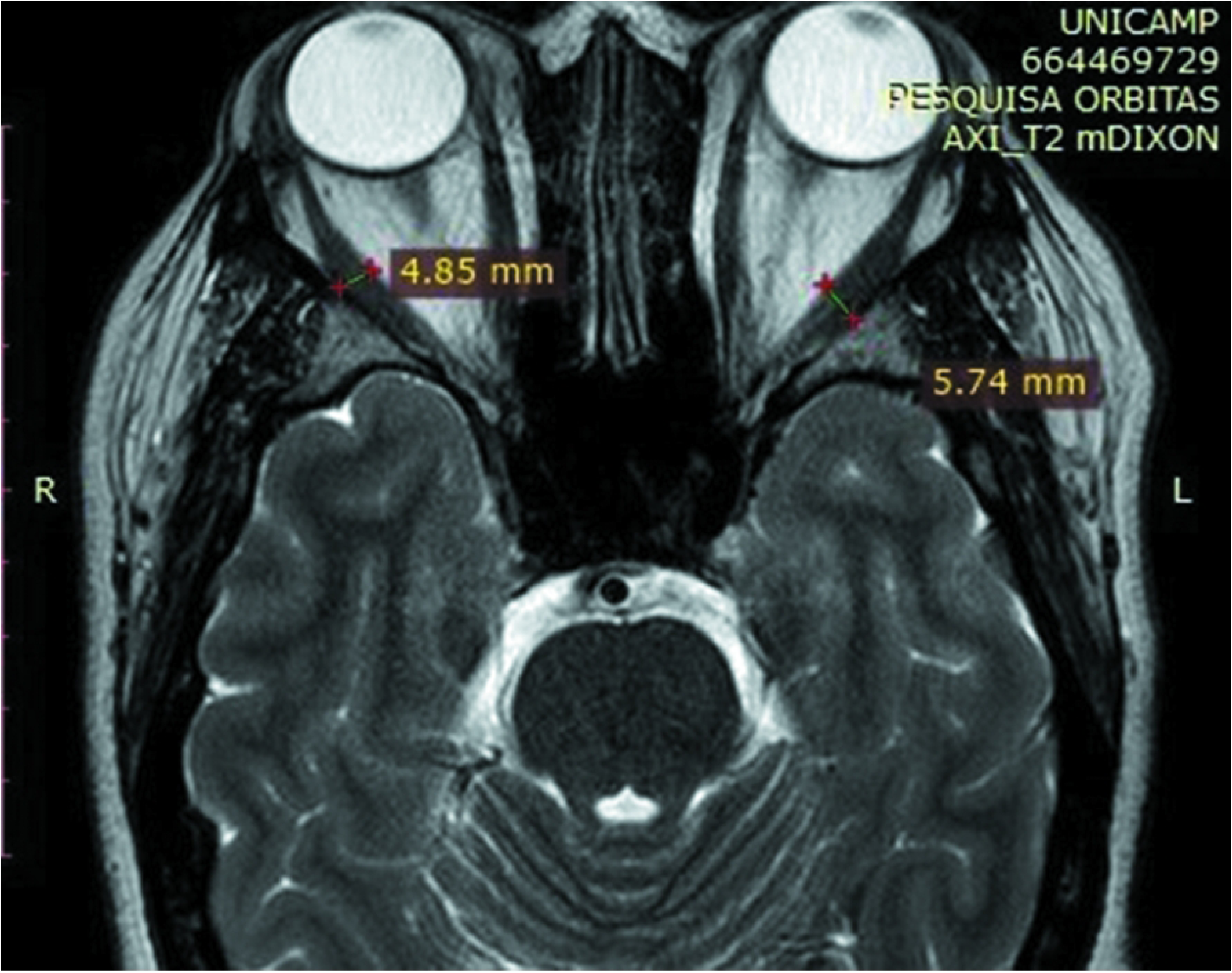
Extraocular muscle measurement technique.

To describe the sample profile according to the variables under study, frequency tables of categorical variables were created with absolute frequency (n) and percentage (%) values. For quantitative variables, descriptive measures were obtained (mean, standard deviation, minimum, median and maximum). The groups were compared using the Chi-square/Fisher’s exact test for categorical variables and the Kruskal-Wallis test for quantitative variables, followed by Dunn’s test, if necessary.

To evaluate the agreement of numerical measurements between evaluators, the intraclass correlation coefficient (ICC) was used with the presentation of a confidence interval, in which ICC values above 0.70 can be considered to have good reliability. For categorical variables, the simple Kappa coefficient was used, where values lower than 0.40 indicate low agreement; between 0.40 and 0.75 indicate satisfactory agreement and above 0.75 indicate excellent agreement (16, 17, 18).

The significance level adopted for the study was 5% and the statistical analysis was performed on the SAS System for Windows, version 9.4 and on the R software, version 4.2.2.

The study was approved by the local ethical committee (CAAE number 92689218.8.0000.5404) and all study procedures and images were done and acquired after the patients Informed Consent Form signature.

## Results

Of the 31 patient images analyzed, 14 individuals were on the control group (group 1), 11 patients on the GD with inactive ophthalmopathy group (group 2) and 6 on the GD with active ophthalmopathy (group 3).

Comparing the groups considering each muscle for each eye, the only group that demonstrated a statistically significant difference was the right superior oblique muscle with a *p-value* of 0.0213, which after a Dunn Test was identified that the group 2 differed from the others. Separating the data by muscle and eye, all others showed a *p-value* higher than 0.3, as shown on Table 1. For this analysis it was considered the mean of the values obtained by the raters on each muscle.

**Table 1.** Comparison among control group, Graves’disease with inactive or active thyroid eye disease - Kruskal-Wallis Test.

| Muscle | Group 1 (N=14)<br>(Mean ± SD) | Group 2 (N=11)<br>(Mean ± SD) | Group 3 (N=6)<br>(Mean ± SD) | *p-value* |
| --- | --- | --- | --- | --- |

|  | (Median (Min-Max)) | (Median (Min-Max)) | (Median (Min-Max)) |  |
| --- | --- | --- | --- | --- |
| Right Inferior | 4.3 ± 0.6 | 4.3 ± 1.1 | 4.7 ± 1.4 | 0.6192 |
| Rectus | 4.4 (3.1-5.5) | 4.0 (3.5-7.5) | 4.0 (3.7-7.2) |  |
| Left Inferior | 4.1 ± 0.5 | 4.3 ± 0.4 | 4.8 ± 1.4 | 0.5578 |
| Rectus | 4.2 (3.2-4.7) | 4.5 (3.6-4.8) | 4.3 (3.7-7.4) |  |
| Right Superior | 3.9 ± 0.7 | 3.9 ± 0.7 | 4.4 ± 1.7 | 0.9990 |
| Rectus | 4.2 (2.3-4.7) | 3.9 (2.7-5.0) | 3.9 (3.2-7.8) |  |
| Left Superior | 4.0 ± 0.7 | 4.1 ± 0.7 | 4.3 ± 1.9 | 0.7817 |
| Rectus | 3.8 (3.2-5.6) | 4.2 (2.6-5.0) | 3.9 (2.7-8.1) |  |
| Right Medial | 4.1 ± 0.6 | 4.3 ± 1.0 | 4.0 ± 1.5 | 0.6123 |
| Rectus | 4.1 (2.8-5.3) | 4.1 (3.2-6.9) | 3.5 (2.8-6.6) |  |
| Left Medial | 4.0 ± 0.6 | 4.0 ± 1.0 | 4.1 ± 1.3 | 0.6428 |
| Rectus | 3.9 (2.9-5.4) | 3.7 (2.9-6.3) | 3.4 (2.9-5.8) |  |
| Right Lateral | 4.2 ± 0.8 | 4.1 ± 0.7 | 4.0 ± 0.6 | 0.7054 |
| Rectus | 4.5 (2.6-5.7) | 4.2 (2.6-5.1) | 3.9 (3.2-5.1) |  |
| Left Lateral | 4.2 ± 0.5 | 4.1 ± 0.6 | 4.4 ± 0.9 | 0.5655 |
| Rectus | 4.3 (3.2-4.9) | 4.0 (3.0-5.3) | 4.4 (3.3-5.9) |  |
| Right Superior | 2.9 ± 0.2 | 2.6 ± 0.3 | 3.0 ± 0.2 | 0.0213 |
| Oblique | 2.9 (2.6-3.2) | 2.5 (2.2-3.1) | 3.0 (2.7-3.2) |  |
| Left Superior | 3.0 ± 0.4 | 2.8 ± 0.4 | 2.8 ± 0.3 | 0.3019 |
| Oblique | 2.9 (2.2-3.8) | 2.7 (2.1-3.5) | 2.9 (2.3-3.1) |  |
| Right Inferior | 2.6 ± 0.4 | 2.4 ± 0.3 | 2.4 ± 0.5 | 0.3227 |
| Oblique | 2.6 (2.0-3.3) | 2.3 (2.0-2.8) | 2.6 (1.5-2.9) |  |
| Left Inferior | 2.5 ± 0.4 | 2.4 ± 0.2 | 2.3 ± 0.5 | 0.5314 |
| Oblique | 2.5 (1.8-3.5) | 2.3 (2.1-2.7) | 2.3 (1.6-3.0) |  |

N, Number of patients; SD, Standard Deviation; Min, Minimum; Max, Maximum. Control group (1), GD with inactive (2) or active (3) TED.

The concordance between raters (Table 2) regarding changed eye structure, was also evaluated considering each of the groups individually to understand if the raters were obtaining similar conclusions in each muscle on each eye. In the control group it was not possible to measure the concordance between raters on the inferior and superior rectus on both eyes and on extra and intraconal fat, however on the medial rectus, lateral rectus, superior oblique in both eyes, as well on the lacrimal gland, the concordance was not statistically considerable since the confidence interval presented negative numbers. The only valid concordance level was the proptosis value with satisfactory concordance.

**Table 2.** Concordance between raters on the enlargement of the eye muscles.

|  | Group 1 (N=14) | Group 2 (N=11) | Group 3 (N=6) |
| --- | --- | --- | --- |
| Muscle | Simple Kappa Coefficient<br>Confidence Interval | Simple Kappa Coefficient<br>Confidence Interval | Simple Kappa Coefficient<br>Confidence Interval |
| Right Inferior Rectus | Unable to measure | 1.0000<br>[1.0000;1.0000] | 1.0000<br>[1.0000;1.0000] |
| Left Inferior Rectus | Unable to measure | Unable to measure | 1.0000<br>[1.0000;1.0000] |
| Right Superior Rectus | Unable to measure | Unable to measure | 1.0000<br>[1.0000;1.0000] |
| Left Superior Rectus | Unable to measure | Unable to measure | 1.0000<br>[1.0000;1.0000] |
| Right Medial Rectus | 0.6316<br>[-0.0153;1.0000] | 0.6207<br>[-0.0352;1.0000] | 1.0000<br>[1.0000;1.0000] |
| Left Medial Rectus | 0.0000<br>[-0.2545;0.0440] | 0.6207<br>[-0.0352;1.0000] | 1.0000<br>[1.0000;1.0000] |
| Right Lateral Rectus | 0.0541<br>[-0.4765;0.5846] | 0.3889<br>[-0.3028;1.0000] | Unable to measure |
| Left Lateral Rectus | 0.1515<br>[-0.4241;0.7271] | 0.6207<br>[-0.0352;1.0000] | 0.5714<br>[-0.1214;1.0000] |
| Right Superior Oblique | 0.0000<br>[-0.4038;-0.0100] | 0.6207<br>[-0.0352;1.0000] | Unable to measure |
| Left Superior Oblique | 0.4324<br>[-0.1001;0.9649] | Unable to measure | Unable to measure |
| Proptosis | 0.5758<br>[0.0504;1.0000] | 0.8136<br>[0.4713;1.0000] | 1.0000<br>[1.0000;1.0000] |
| Extraconal Fat | Unable to measure | 0.1538<br>[-0.0894;0.3971] | Unable to measure |
| Intraconal Fat | Unable to measure | 0.2143<br>[-0.1657;0.5942] | 0.0000<br>[-0.7014;0.1300] |
| Lacrimal Gland | 0.2222<br>[-0.0735;0.5179] | 0.0000<br>[-0.3347;0.0588] | 0.0000<br>[-0.4716;0.0716] |

N, Number of patients.

In GD and inactive ophthalmopathy group the right inferior rectus presented perfect concordance; the left inferior rectus, superior rectus on both eyes and left superior oblique concordance was not able to be measured. Similar as in the control group, the medial rectus on both eyes, lateral rectus on both eyes, right superior oblique, proptosis, intraconal fat, extraconal fat and lacrimal gland were not statistically considerable due to the confidence interval presenting negative numbers.

In GD with active ophthalmopathy group, the inferior rectus, superior rectus and medial rectus, all on both eyes, presented perfect concordance along with the proptosis. Right lateral rectus, superior oblique on both eyes and extraconal fat were not able to be measured, while the left lateral rectus, intraconal fat and lacrimal gland presented negative numbers on the confidence interval.

Concordance between the values measured by the raters was evaluated to verify if the results obtained were similar, since they both were using the same technique to obtain these values for each eye muscle, as shown on Table 3.

**Table 3.** Descriptive measures of the eye muscles and concordance between values obtained by each rater.

| Muscle | Group 1 (N=14) |  | Group 2 (N=11) |  | Group 3 (N=6) |  |
| --- | --- | --- | --- | --- | --- | --- |
|  | Rater 1 | ICC (IC95%) | Rater 1 | ICC (IC95%) | Rater 1 | ICC (IC95%) |
|  | Rater 2 |  | Rater 2 |  | Rater 2 |  |
| Right Inferior | 4.17 |  | 4.11 |  | 4.52 |  |
| Rectus | 4.51 | 0.40 (0.09, 0.81) | 4.54 | 0.60 (0.23, 0.88) | 4.88 | 0.74 (0.30, 0.95) |
| Left Inferior | 3.92 |  | 4.27 |  | 4.65 |  |
| Rectus | 4.29 | 0.27 (0.03, 0.81) | 4.37 | 0.00 | 4.92 | 0.85 (0.50, 0.97) |
| Right Superior | 4.09 |  | 3.71 |  | 4.07 |  |
| Rectus | 3.78 | 0.36 (0.07, 0.80) | 4.19 | 0.00 | 4.78 | 0.81 (0.42, 0.96) |
| Left Superior | 4.13 |  | 3.88 |  | 3.89 |  |
| Rectus | 3.96 | 0.19 (0.01, 0.87) | 4.33 | 0.08 (0.00, 1.00) | 4.80 | 0.78 (0.36, 0.96) |
| Right Medial | 4.04 |  | 4.28 |  | 3.75 |  |
| Rectus | 4.18 | 0.68 (0.37, 0.89) | 4.35 | 0.35 (0.05, 0.84) | 4.27 | 0.87 (0.56, 0.97) |
| Left Medial | 3.95 |  | 3.88 |  | 3.61 |  |
| Rectus | 4.10 | 0.28 (0.04, 0.81) | 4.21 | 0.51 (0.15, 0.86) | 4.50 | 0.51 (0.09, 0.92) |
| Right Lateral | 4.26 |  | 3.99 |  | 3.89 |  |
| Rectus | 4.14 | 0.40 (0.10, 0.81) | 4.15 | 0.49 (0.14, 0.85) | 4.07 | 0.42 (0.04, 0.92) |
| Left Lateral | 4.07 |  | 3.81 |  | 4.23 |  |
| Rectus | 4.39 | 0.00 | 4.38 | 0.00 | 4.65 | 0.48 (0.07, 0.92) |
| Right Superior | 2.78 |  | 2.28 |  | 2.87 |  |
| Oblique | 3.11 | 0.00 | 3.12 | 0.00 | 3.07 | 0.00 |
| Left Superior | 2.89 |  | 2.52 |  | 2.57 |  |
| Oblique | 3.11 | 0.33 (0.06, 0.80) | 3.25 | 0.03 (0.00, 1.00) | 3.07 | 0.00 |

N, Number of patients; ICC, Intraclass Correlation Coefficient; IC, Interval of Confidence.

On the control and GD with inactive ophthalmopathy groups none of the muscles presented a good reliability intraclass correlation coefficient (ICC), being the closest ones the right medial rectus on the control group with an ICC of 0.68 and the right inferior rectus on the GD with inactive ophthalmopathy, with an ICC of 0.60.

The GD with active ophthalmopathy, the right and left inferior rectus, right and left superior rectus, and the right medial rectus presented a good reliability ICC with values of 0.74, 0.85, 0.81, 0.78 and 0.87 respectively. The other muscles had values inferior to 0.51.

The lacrimal gland changes were evaluated separately between groups by each rater to confirm the reliability of the measures for each rater. Only rater 1 had a statistically significant result with a p-value of 0.0345, as seen on Table 4.

**Table 4.** Comparison of the augmented lacrimal glands between groups by each rater.

|  | Group 1 (N=14) | Group 2 (N=11) | Group 3 (N=6) | Total (N=31) |  |
| --- | --- | --- | --- | --- | --- |
| Rater | No (N (%)) | No (N (%)) | No (N (%)) | No (N (%)) | *p-value* |
|  | Yes (N (%)) | Yes (N (%)) | Yes (N (%)) | Yes (N (%)) |  |
| 1 | 8 (57.1%) | 1 (9.1%) | 1 (16.7%) | 10 (32.3%) | 0.0345 |
|  | 6 (42.9%) | 10 (90.9%) | 5 (83.3%) | 21 (67.7%) |  |
| 2 | 2 (14.3%) | 2 (18.2%) | 1 (16.7%) | 5 (16.1%) | 1.0000 |
|  | 12 (85.7%) | 9 (81.8%) | 5 (83.3%) | 26 (83.9%) |  |
| N, Number of patients. |  |  |  |  |  |

On Table 5 the comparison between groups was made separated by rater, and like on table 1, the only muscle with a statistically significant difference was the right superior oblique muscle with a *p-value* of 0.0185. All other muscles present a *p-value* higher than 0.2.

**Table 5.**
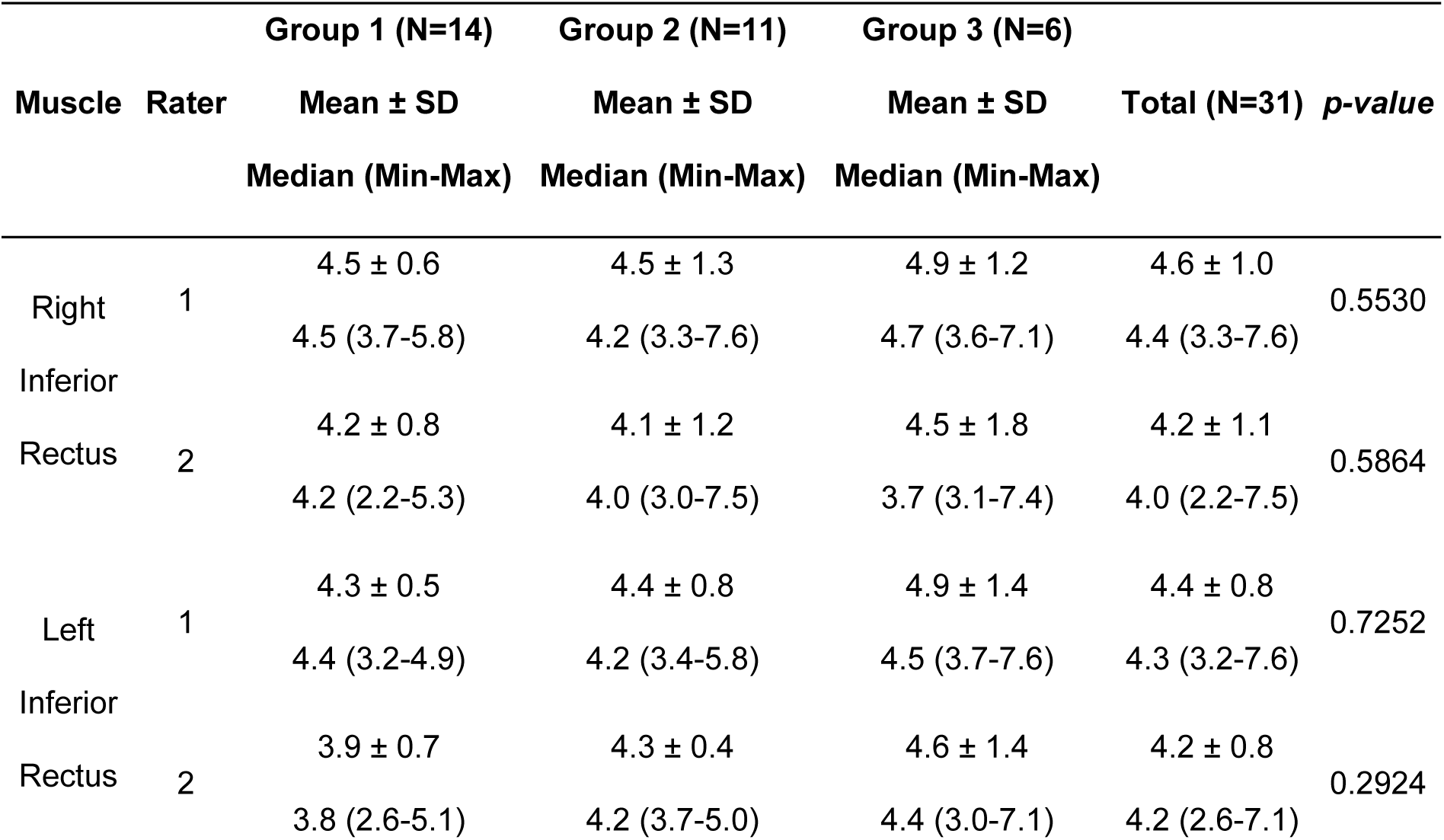
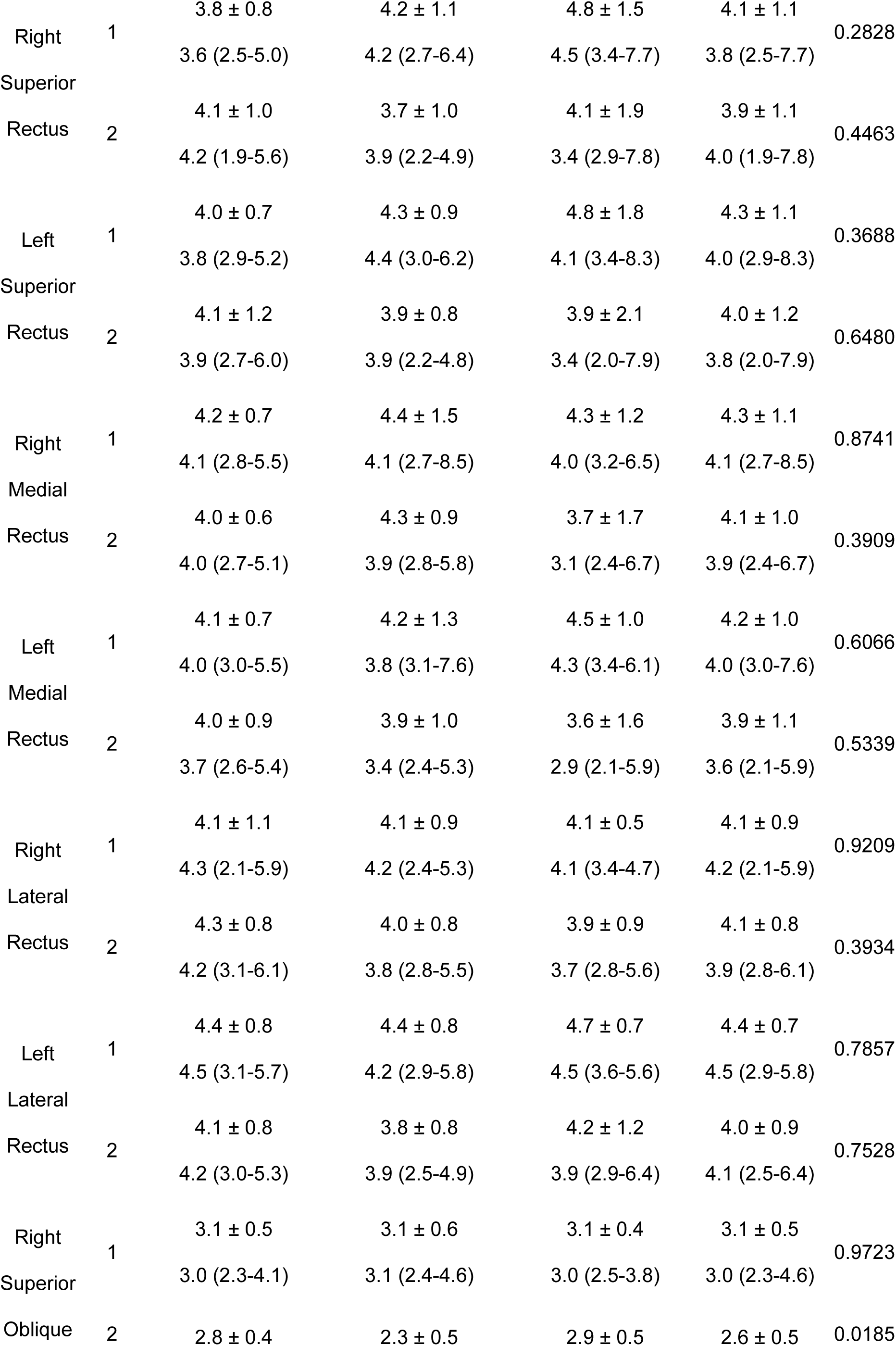
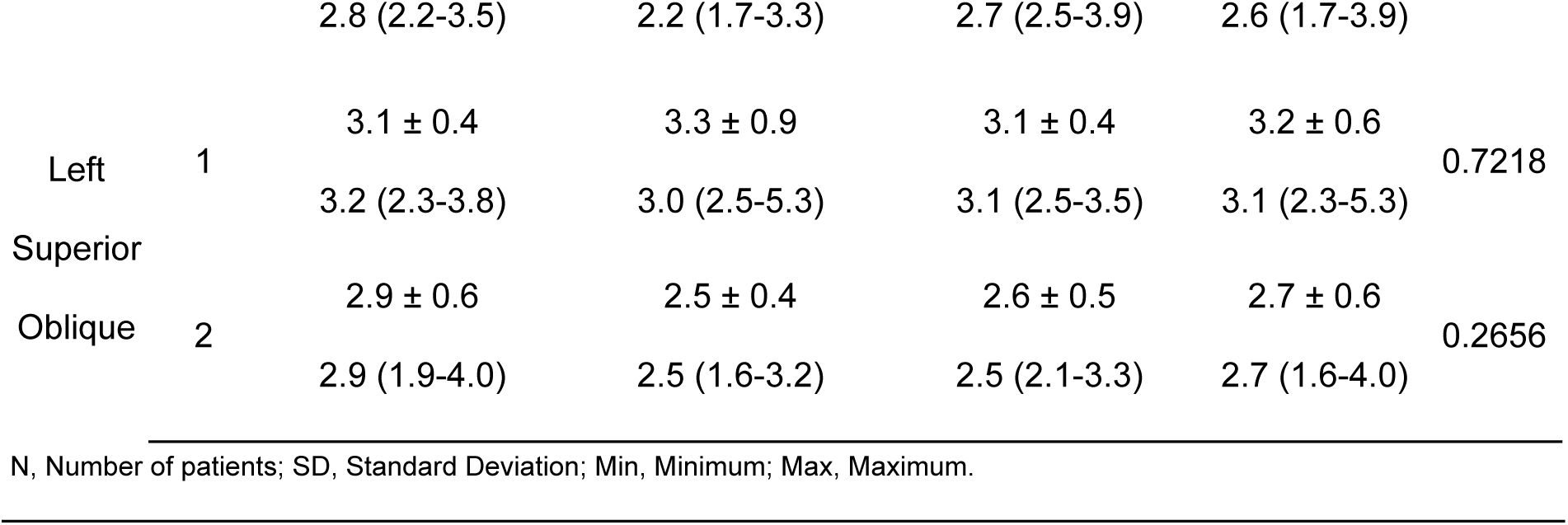
Comparison between groups separated by rater - Kruskal-Wallis Test.

## Discussion

Measuring extraocular muscle thickness in thyroid eye disease can reveal important information regarding inflammatory activity, severity and progression of the disease, being one of the radiological parameters most investigated. However, taking these measurements properly can be a challenge because, despite the current guidance on the methods, it will highly depend on the rater’s knowledge and experience performing this task (19).

In the present study, when considering the median of the values obtained by the raters in each muscle evaluated, there is no difference between the muscles, with the exception of the right superior oblique, with GD and inactive ophthalmopathy differing from the others. However, there would be a bias in the interpretation of the results since the inactive ophthalmopathy could exhibit muscle measurements similar to the individuals of the control group. When evaluating the values obtained separately by each rater, the rater 1 measurements showed no difference between any of the groups in any muscle, while rater 2 measurements behaved the same way as the median values; indicating that this result alone may have influenced directly on the results obtained with the median values of the raters.

The concordance test between raters considered not the measurements *per se* but if on the raters’ assessment there was an enlargement of the eye structures, not only accounting the eye muscles but also intra and extraconal fat, proptosis and lacrimal gland. Some concordances were not possible to be measured due to one of the raters indicating that there were no changes in any of the patients for that particular eye structure, while others calculated showed a confidence interval with negative numbers, invalidating the significance of the simple kappa coefficient. The only significant value found by the analysis was the proptosis evaluated on the group with GD and inactive ophthalmopathy with a high coefficient of 80% and a perfect concordance on the right inferior rectus. On the group with GD and active ophthalmopathy there was perfect concordance on the inferior, superior and medial rectus on both eyes, as well on proptosis, however this level of concordance can be directly attributed to the low number of patients included on the analysis, only six.

Looking at the whole scenario, the concordance values were increasing the statistical significance as the number of patients in each group were decreasing, which can possibly indicate that the variability can be an ally in obtaining reliable statistical results.

When evaluating the concordance between methods and its descriptive measures, control group and GD with inactive TED did not present a good reliability intraclass correlation coefficient on any extraocular muscle, however GD with active TED presented it on inferior and superior rectus, on both eyes, and on the right medial rectus. These results suggest that the most affected structures, as expected in TED with inflammatory activity, are obviously easier to recognize as altered, a fact that may not be as evident when analyzing images of individuals without TED or with TED in an inactive phase of inflammation.

The lacrimal gland was once appointed by several studies as one of the most affected by TED (20) with changes easier to detect on imaging analysis since the normal size of the gland is more well-established than the extraocular muscles (21). Considering this fact, a comparison was made between groups and raters not by measurements but determining if the gland size was increased and only rater 1 found differences between groups, unlike rater 2.

On an overview of all results and considering the real medical practice on assessing eye structure changes, we could identify differences on the results for each rater. One rater had more reliable results evaluating eye muscles, while the other had better results assessing other eye structures, which can be due to the experience with the disease and with the imaging analysis, two factors impacting directly on the quality and reliability of the method. Another factor that can affect the evaluation and is not rater-related is the image quality and definition; this definition can directly interfere with the enlargement measurement, since the line used to obtain this metric is placed from one edge of the muscle to another, and the line position may differ depending on the image definition. To achieve a proper image quality for the evaluation, the recommendation is the use of an MRI machine with a magnetic field of 3T or higher.

## Conclusion and Future Directions

This study presented difficulties that impacted directly on its proposal, one of them being the low number of patients available and availability to perform MRI. The study showed that the method of measuring can help on diagnosis but it is far to be a gold standard on evaluating a TED since each rater, despite using the same images, can be performing the measurements on their own way, for example, not using the same image spot to determine muscle thickness or having different parameters of normality, even more for extraocular muscles. More raters would also help to identify the variability of data obtention between raters to determine their differences when executing the measurements. On the other hand, a strong point was the evaluation of the images by two experienced radiologists in the area, a fact that increases the reliability of the findings. Furthermore, another strength was based on the evaluation of the groups according to the presence of TED and inflammatory activity and comparison with control group subjects without evidence of TED.

We concluded, with the available set of data, that the method is not reliable as a standalone method for evaluation while there is no gold standard on how to perform it and must continue to be used as supplementary information to corroborate clinical and laboratorial findings. A combination of different methods such as image and laboratory findings and/or clinical findings can result in a more elaborate and reliable evaluation of the patient’s condition. Additional studies are needed to establish a good practice for imaging on TED; its importance is already determined but its use needs a guideline to facilitate disease diagnosis and patient follow-up with non-invasive methods and available tools that nowadays are standard on hospitals with imaging resources.

## Data Availability

All relevant data are within the manuscript and its Supporting Information files.

## Acknowledgements

We acknowledge all patients who voluntarily participated in our study.

We thank the National Council of Technological and Scientific Development (CNPq) for the scholarship 303068/2021-3 resource.

We thank Cleide Aparecida Moreira Silva and Marcelo Tavares from the statistical service of the School of Medical Science of University of Campinas.

## References

1. Riguetto CM, Neto AM, Tambascia MA, Zantut-Wittmann DE. 2019. The relationship between quality of life, cognition, and thyroid status in Graves’ disease. Endocrine. 63(1):87–93. doi: 10.1007/s12020-018-1733-y.

2. Łacheta D, Miśkiewicz P, Głuszko A, Nowicka G, Struga M, Kantor I, Poślednik KB, Mirza S, Szczepański MJ. 2019. Immunological aspects of Graves’ ophthalmopathy. Biomed Res Int [Internet]. [cited 2025 Aug 23];2019:7453260. doi: 10.1155/2019/7453260.

3. Fox TJ, Anastasopoulou C. 2022. Graves orbitopathy. In: StatPearls [Internet]. Treasure Island (FL): StatPearls Publishing; [updated 2022 Aug 29; cited 2025 Aug 23]. Available from: https://www.ncbi.nlm.nih.gov/books/NBK549889/.

4. Ozgen A, Ariyurek M. 1998. Normative measurements of orbital structures using CT. AJR Am J Roentgenol. 170(4):1093–6. doi: 10.2214/ajr.170.4.9530066.

5. Wolf A, Jaafar M, Olsen C, Kadom N. 2008. Normative values for pediatric extraocular muscle position. Invest Ophthalmol Vis Sci. 49(13):1805.

6. Zhang ZH, Chen Y, Wang Y, Meng W, Fang HY, Xu DD, Jin ZY.. 2013. Normative measurements of extraocular musculature by multislice computed tomography. Chin Med Sci J. 27(4):232–6. doi: 10.1016/s1001-9294(13)60007-3.

7. Rana K, Juniat V, Rayan A, Patel S, Selva D. 2022. Normative measurements of orbital structures by magnetic resonance imaging. Int Ophthalmol. 42(12):3869–75. doi: 10.1007/s10792-022-02407-1.

8. Maurya RP, Ananya PR, Kadir SMU, Singh VP, Das D, Gupta S, Agrawal S, Singh V, Roy M. 2021. Recent advances in thyroid eye disease: an overview. IP Int J Ocul Oncol Oculoplasty. 7(2):117–30. doi: 10.18231/j.ijooo.2021.027.

9. Su Y, Liu X, Fang S, Huang Y, Li Y, Zhong S, Wang Y, Zhang S, Zhou H, Sun J, Fan X. 2022. Age-related difference in extraocular muscles and its relation to clinical manifestations in an ethnically homogenous group of patients with Graves’ orbitopathy. Graefes Arch Clin Exp Ophthalmol. 260(2):583–9. doi: 10.1007/s00417-021-05377-9.

10. North VS, Freitag SK. 2019. A review of imaging modalities in thyroid-associated orbitopathy. Int Ophthalmol Clin. 59(4):81–93. doi: 10.1097/IIO.0000000000000289.

11. Klingenstein A, Samel C, Garip-Kübler A, Hintschich C, Muller-Lisse UG. 2022. Cross-sectional computed tomography assessment of exophthalmos in comparison to clinical measurement via Hertel exophthalmometry. Sci Rep. 12:11973. doi: 10.1038/s41598-022-16131-4.

12. Wnuk E, Maj E, Jabłońska-Pawlak A, Jeczeń M, Rowińska-Berman K, Rowiński O. 2022. Validation of exophthalmos magnetic resonance imaging measurements in patients with Graves’ orbitopathy, compared to ophthalmometry results. Pol J Radiol. 87:e539–e544. doi: 10.5114/pjr.2022.119939.

13. Cevik Y, Taylan Sekeroglu H, Ozgen B, Erkan Turan K, Sanac AS. 2021. Clinical and radiological findings in patients with newly diagnosed Graves’ ophthalmopathy. Int J Endocrinol [Internet]. [cited 2025 Aug 23];2021:5513008. doi: 10.1155/2021/5513008.

14. Hutchings KR, Fritzhand SJ, Esmaeli B, Koka K, Zhao J, Ahmed S, Debnam JM. 2023. Graves’ eye disease: clinical and radiological diagnosis. Biomedicines. 11(2):312. doi: 10.3390/biomedicines11020312.

15. Xu L, Li L, Xie C, Guan M, Xue Y. 2017. Thickness of extraocular muscle and orbital fat in MRI predicts response to glucocorticoid therapy in Graves’ ophthalmopathy. Int J Endocrinol [Internet]. [cited 2025 Aug 23];2017:3196059. doi: 10.1155/2017/3196059.

16. Conover WJ. 1999. Practical nonparametric statistics. 3rd ed. New York (NY): John Wiley & Sons.

17. Fleiss JL. 1981. Statistical methods for rates and proportions. 2nd ed. New York (NY): John Wiley & Sons.

18. Streiner DL, Norman GR. 1995. Health measurement scales: a practical guide to their development and use. 2nd ed. New York (NY): Oxford University Press.

19. Lee JY, Bae K, Park KA, Lyu IJ, Oh SY. 2016. Correlation between extraocular muscle size measured by computed tomography and the vertical angle of deviation in thyroid eye disease. PLoS One. 11(1):e0148167. doi: 10.1371/journal.pone.0148167.

20. Luccas R, Riguetto CM, Alves M, Zantut-Wittmann DE, Reis F. 2024. Computed tomography and magnetic resonance imaging approaches to Graves’ ophthalmopathy: a narrative review. Front Endocrinol (Lausanne). 14:1277961. doi: 10.3389/fendo.2023.1277961.

21. Dalvi VN, Ambhore A, Dhok AP, Onkar PM, Mitra K. 2023. Normal dimensions of the lacrimal gland on magnetic resonance imaging in Indian adult population: a retrospective study. Pan Afr Med J. 45:71. doi: 10.11604/pamj.2023.45.71.38213.

